# Prenatal exposure to polycyclic aromatic hydrocarbons and birth outcomes in a pregnancy cohort in Nairobi, Kenya

**DOI:** 10.1101/2025.08.01.25332639

**Authors:** Allison R. Sherris, Priscillah Wanini Edemba, Anne M. Riederer, Judy Adhiambo, Lewis Olweywe, Prestone Owiti, Catherine J. Karr, John Kinuthia, Barbra A. Richardson, Christopher D. Simpson, Christopher Zuidema, Elizabeth Maleche-Obimbo, Sarah Benki-Nugent

## Abstract

Prenatal exposure to polycyclic aromatic hydrocarbons (PAHs) has been linked to lower birth weight (BW) and shorter gestational age (GA) at delivery. Most research has focused on populations in high-income countries, leaving low- and middle-income countries (LMICs) understudied. We examined associations between pregnancy urinary PAH metabolites and birth outcomes in a prospective cohort in Nairobi, Kenya.

The study population was drawn from women and their newborn babies enrolled in a pregnancy cohort in Nairobi, Kenya. Third-trimester urinary mono-hydroxylated PAH metabolites (OH-PAH) were measured and infant BW and GA were ascertained using medical record abstraction and self-report; BW-for-GA z-scores were computed. Linear and modified Poisson regression models were used to estimate the effects of five OH-PAHs on birth outcomes; Weighted Quantile Sum regression was used to explore OH-PAH mixture effects.

Among 353 mother-infant pairs, the median infant BW was 3.2 kg and10.7% were born preterm. All OH-PAH metabolites were present in urine of >99% of mothers. Individual OH-PAH concentrations were not associated with BW or BW-for-GA z-scores. One metabolite, 2-hydroxyphenanthrene, was associated with shortened gestation (RR = -1.6 day per doubling in concentration, 95% confidence interval: -3.1, - 0.1). This association was attenuated after adjusting for self-reported exposure to household fuel and outdoor combustion and was strengthened among female infants, but not male infants, in sex-stratified analyses. Other metabolites and the OH-PAH mixture were not associated with birth outcomes.

Our findings suggest that exposure to 2-hydroxyphenanthrene may have a modest adverse effect on pregnancy duration, with potential sex-specific differences in association. No associations were observed for markers of fetal growth. These results highlight need for further studies on sex-specific vulnerabilities and the role of environmental co-exposures in impacting birth outcomes in LMICs.

## 1. Introduction

Polycyclic aromatic hydrocarbons (PAHs) are a class of air pollutants originating from the burning of organic materials including biomass, fossil fuels, and tobacco products, as well as charred foods.^1^ Exposure to PAHs may occur through inhalation of gaseous PAHs or those bound to ambient particulate matter (PM), ingestion of grilled or charred food, and contact with polluted surfaces.^2–4^ PAHs are categorized into high molecular weight (HMW) and low molecular weight (LMW) compounds, which have different metabolization and biological impacts. Exposure to both classes of PAHs is well-documented in the general population and among pregnant individuals in the United States,^5–7^ though fewer studies have characterized exposure in low- and middle-income countries (LMICs). The health effects of PAHs are of concern in LMICs where household combustion of solid and liquid cooking fuels, waste burning, traffic emissions, waste burning, and industrial activities can contribute to high levels of household and ambient air pollution.^8–11^

PAH exposure is linked to a variety of adverse health outcomes including respiratory illness, cardiovascular disease, and cancer, with most studies focused on HMW PAHs such as benzo[*a*]pyrene.^12–15^ Recent literature has also explored the reproductive health effects of PAHs, though findings have been inconsistent.^16–18^ While a recent large U.S.-based cohort study found that certain urinary metabolites of LMW PAHs were significantly associated with reduced gestational age (GA) at delivery,^19^ only three out of ten studies included in a recent review identified similar adverse associations.^17^ Two recent meta-analyses also found no association between one maternal urinary LMW PAH metabolite, 1- hydroxypyrene (1-OH-PYR), and birth weight (BW) among five included studies.^16,17^ However, meta-analyses focused on exposure to airborne HMW PAHs and dietary sources of PAHs have found stronger adverse associations with BW.^16,18^ These potential impacts have been attributed to factors including oxidative stress, systemic inflammation, endocrine disruption, and other epigenetic impacts, some of which have been found to be modified by fetal sex.^20,21^ Prior epidemiologic studies have used a variety of study designs, outcome measures, and exposure assessment methods and have primarily been conducted in high-income countries, as well as eastern Europe and China. Indeed, none of the aforementioned reviews included studies from Africa, Latin America, or Oceania.^16–18^ As communities in LMICs likely experience different PAH exposure levels, sources, and co-exposures, this is an important gap in understanding of global PAH health effects.

Sub-Saharan Africa (SSA) is a region with documented high levels of environmental PAHs and ambient air pollution, along with a scarcity of epidemiologic research on related health impacts.^10,22,23^ This region also faces high rates of adverse birth outcomes. In 2020, Over 10% of infants were born preterm in SSA compared to under 8% Europe , and approximately 14% were born with low birth weight (<2500 g) compared to 7.6% in Europe.^24,25^ Preterm delivery is the leading cause of neonatal morbidity worldwide, and infants born preterm or low birth weight are at higher risk of adverse health outcomes, including neurodevelopmental and respiratory outcomes.^26–28^ Given the combined burden of both environmental exposures and adverse birth outcomes, there is a critical need for research on the health impacts of air pollution and PAHs in SSA.

This study evaluates associations between prenatal LMW PAH exposure and birth outcomes in a prospective birth cohort in Nairobi, Kenya. We leverage measurements of prenatal urinary PAH metabolite concentrations to determine the impact of individual metabolites and metabolite mixtures on BW (BW), gestational age (GA), preterm birth, and birth weight for gestational age (BW-GA) z-scores. We also evaluate whether associations are modified by child sex. Together, these findings address a critical knowledge gap regarding the perinatal health effects of LMW PAH exposure in Sub-Saharan Africa.

## 2. Materials and Methods

### 2.1 Study population and inclusion criteria

This study population included pregnant women and their infants from the Air Pollution Exposures in Early Life and Brain Development in Children (ABC) Study.^29^ In brief, pregnant women age 18-40 years were recruited and enrolled from January 10, 2022 to November 29, 2022 during antenatal visits at the Dandora 2 Health Centre from 2021-2023. The Dandora neighborhoods of Nairobi are adjacent to a large dump site with routine waste burning. Inclusion criteria were residence in Nairobi and planning to reside in the same residence for at least one year. Participants with severe pregnancy complications were excluded. At enrollment, participants completed questionnaires describing sociodemographic characteristics and household, occupational, and outdoor environmental exposures.

Inclusion criteria for the present analysis were: 1) live singleton birth, 2) data on BW and GA, and 3) urinary PAH metabolite and specific gravity (SG) measurements in the third trimester of pregnancy. In sensitivity analyses, we excluded participants with evidence of prenatal smoking (based on maternal self- report of smoking in the 24-hours prior to urine collection and/or maternal urinary cotinine concentration above 200 ng/mL during pregnancy) and, in separate models, positive HIV status.

### 2.2 Ethics Statement

All research activities were approved by the Kenyatta National Hospital (KNH) - University of Nairobi Ethics and Research Committee and the University of Washington Human Subjects Division Institutional Review Board. All participants provided prior informed consent.

### 2.3 Urinary sample collection and laboratory procedures

Many low-molecular weight (LMW) PAHs are metabolized to monohydroxylated metabolites (OH- PAHs). OH-PAHs are commonly detected in urine in the general population and during pregnancy.^5^ Spot urine samples were collected from ABC participants at the third trimester visit and the enrollment visit (if enrollment was earlier than the third trimester) for analysis of OH-PAHs. Urine was transported to the clinic laboratory at KNH where it was stored at -70°C then shipped frozen to the University of Washington (Seattle, USA) where it was extracted and analyzed for seven OH-PAH metabolites, including 1-napthol (1-OH-NAP), 2-OH-napthol (2-OH-NAP), 2-hydroxyfluorene (2-OH-FLUO), 9- hydroxyfluorene (9-OH-FLUO), 2-hydroxyphenanthrene (2-OH-PHEN), 3-hydroxyphenanthrene (3-OH- PHEN), and 1-hydroxypyrene (1-OH-PYR) using high performance liquid chromatography (HPLC) with fluorescence detection based on the method reported by Chetiyanukornkul et al.^30^, with modifications. 2- OH-FLUO and 9-OH-FLUO co-elute under the HPLC conditions used, so a single value is reported for the sum of these two metabolites (2/9-OH-FLUO). The HPLC system was an Agilent 1100 series and the column was an Agilent Poroshell 120 SB-C18 (100 × 2.1 mm, 2.7 μm). Mobile phases were 10 mM sodium acetate (pH = 5) and methanol. In brief, 10 ml urine was amended with 0.1 mL of ascorbicacid solution and 5 mL of acetate buffer (pH = 5.5) and allowed to incubate at 37 °C for three hours in a shaking water bath. Next, 1.6 mL of methanol was added to each sample and the samples were placed in solid phase extraction columns (Supelclean LC-18 500g; Sigma-Aldrich, St. Louis, MO USA). The columns were rinsed with 5mL of 40:60 methanol:water (v:v), then eluted with 5mL of 80:20 methanol:water (v:v). Each sample was reduced in volume to approximately 2 mL in a TurboVap evaporator (Biotage, San Jose, CA USA) at 55°C. Samples were then mixed with 4.5 mL acetonitrile and allowed to evaporate to a final volume of approximately 1 mL. This step was then repeated a second time to remove all residual water. The samples were next vortexed with 0.02 mL dimethylsulfoxide, allowed to evaporate to near dryness, and then reconstituted in 0.2 mL of a 40:60 methanol:water (v:v). The samples were again vortexed and sonicated for five minutes each before filtration into HPLC vials for analysis. OH-PAH metabolites were identified by matching analyte peaks with those of a series of authentic standards, and quantified based on calibration curves prepared from authentic standards.

Limits of detection (LOD) were determined based on the lowest detectable calibration solution and were as follows: 1-OH-NAP: 3.52 ng/mL; 2-OH-NAP: 0.035 ng/mL; 2/9-OH-FLUO: 0.013 ng/mL; 2-OH-PHEN: 0.002 ng/mL; 3-OH-PHEN: 0.002 ng/mL; 1-OH-PYR 0.011 ng/mL. No OH-PAH metabolites were present above the LOD in assay blanks. Assay precision determined from replicate analyses of laboratory water samples that were spiked with OH-PAHs and processed in an identical manner to the urine samples ranged between +/- 21% for 2-OH-NAP to +/- 42% for 2/9-OH-FLUO. Values below the detection limit were substituted with LOD/√2.

Cotinine was measured in urine at the UW Seattle laboratory using 10 ng/mL and 200 ng/mL Cotinine Rapid Test Cassettes (Acro Biotech Inc., Montclair, CA, USA). Lateral flow immunoassay test strips such as these have been shown to be sensitive, specific, and accurate relative to the gold standard liquid-chromatography tandem mass spectrometry method.^31^ Although a participant’s smoking status cannot be definitively determined based on cotinine levels in a single urine sample,^31^ > 200 ng/mL, a commonly used cut-off for identifying active tobacco smoking,^32^ and 10 – 200 ng/mL to indicate potential environmental tobacco smoke (ETS) exposure were used based on our own observations in a multi-site study of 1,603 U.S. pregnant non-smokers.^33^ Participants were categorized into three urinary cotinine categories, < 10 ng/mL, 10-200 ng/mL, and > 200 ng/mL to indicate levels of exposure to ETS.

SG was measured using a handheld refractometer at the clinic laboratory at KNH. Thirteen samples initially measured using a paper strip were remeasured with the refractometer at a later date (after a freeze-thaw cycle); their mean (SD) specific gravity was slightly higher than the rest of the samples—1.021 (0.009) vs. 1.014 (0.006), respectively.

### 2.4 Self-reported environmental pollutant exposures

Questionnaire-based data on exposure to combustion sources was included to control for confounding by co-pollutants generated along with PAHs. At enrollment, participants were asked about indoor exposure to smoke from cooking fuels including wood, kerosene, charcoal, and brickettes. Exposure was evaluated as *daily, most days, some days, rarely* or *not at all*. A three-level variable was derived for *indoor pollutant cooking fuel exposure* defined as exposure to smoke from wood, kerosene, charcoal, or brickettes *daily or most days*, *some days*, or *rarely or not at all*.

Enrollment questionnaires also collected information on household indoor exposure to other combustion sources including incense, kerosene lamps, mosquito repellent, cigarette smoke, marijuana smoke, candles, and rubbish burning; outdoor household exposure to their own or neighbors’ cooking smoke; outdoor exposure (beyond the home) and work-related exposure to cooking smoke, vehicle smoke, factory smoke, dumpsite, and rubbish burning; and work-related exposure to marijuana and cigarette smoke.

### 2.5 Pregnancy Outcomes

The primary outcomes for this analysis were BW (kg) and GA (days). Secondary outcomes were BW-GA z-scores and preterm birth (delivery < 37 weeks of gestation). Information on the newborn at delivery including BW, infant sex, and date of birth was collected at Dandora 2 Health Centre for women who gave birth at the study facility and by telephone for those who delivered elsewhere. Data on maternal pregnancy complications and GA at birth were abstracted from medical records (e.g., the Kenya Mother & Child Health Handbook) at the next scheduled study visit. BW-GA z-scores were calculated based on child sex, BW, and GA using the INTERGROWTH-21 standards, which are based on a global reference population that included Kenyan newborns.^34^

GA was initially calculated as the days between reported last menstrual period (LMP) at enrollment and date of birth. Calculated GA was compared to reported GA from medical records (in weeks) for quality assurance as follows. If the participant was missing LMP, reported GA was substituted (N=1). If the calculated GA was available but reported GA was missing and not available for comparison, the participant was excluded (N=2). If the calculated GA was > 42 weeks and reported GA was ≤ 42 weeks (N=30), the reported GA was substituted. If the calculated and reported GA differed by 2 or more weeks and reported GA resulted in a more conservative BW-GA z-score (i.e., closer to 0), reported GA was substituted (N = 39).

### 2.6 Statistical analysis

For descriptive statistics and mixtures methods, OH-PAH concentrations were adjusted for specific gravity (SG) in order to account for urinary dilution as described in Boeniger et al.^35^ To estimate overall OH-PAH exposure for descriptive statistics, we calculated the molar sum of SG-adjusted OH-PAH metabolites In analytic models, we used unadjusted OH-PAH concentrations with model adjustment for urinary SG.

We used linear regression to estimate the risk ratios and 95% confidence intervals for BW, GA, and BW- GA z-scores associated with prenatal exposure to individual OH-PAHs. Modified Poisson regression with robust standard errors was used to evaluate associations with preterm birth. Unadjusted OH-PAH concentrations were log-transformed with base-2; reported outcomes are associated with a doubling in OH-PAH levels.

We performed separate regression models for individual urinary OH-PAHs. Covariates were selected *a priori* if they were 1) confounders directly or indirectly associated with prenatal PAH exposure and birth outcomes or 2) precision variables associated with birth outcomes alone. Models were adjusted for potential confounders in a staged approach. ‘Primary’ models included urinary SG, maternal age (years), maternal employment (homemaker, waged employment, or not employed), maternal education at enrollment (no formal school, pre-primary school, primary school, secondary school, vocational school, or college/university), maternal marital status (divorced/separated/widowed, married, or never married/single), parity at enrollment (0, 1, 2, or 3 or more), and cotinine ≥ 10 ng/mL.

‘Co-exposure’ models captured exposure to combustion sources that might be determinants of PAH exposure and produce co-pollutants that independently impact birth outcomes. These included exposure to dirty cooking fuel, defined above, and exposure to other combustion sources indoors, outdoors, and related to work or daily activities. We performed Principal Components Analysis (PCA) to summarize participant exposure to combustion sources with at least 10% of participants reporting exposure “rarely”, “some days”, “most days” or “daily” (Table S1). PCA yielded seven principal components (PCs) with eigenvalues of > 1 explaining 66% of the variance, with loadings that were dominated by 1) outdoor and occupational dumpsite and rubbish burning exposure; 2) outdoor exposure to neighbor’s cooking smoke and occupational exposure to cigarettes/marijuana; 3) outdoor and occupational vehicle smoke exposure; 4) occupational exposure to kerosene, charcoal, and wood smoke; 5) outdoor and occupational welding smoke exposure; 6) own cooking smoke outdoors; and 7) indoor exposure to incense and mosquito repellent (Table S2). The co-exposure model included these loadings as covariates in addition to dirty cooking fuel.

We explored eefect modification by child sex by using multiplicative interaction terms between OH-PAH concentrations and child sex. We used weighted quantile sum (WQS) regression to explore associations between OH-PAH mixtures and birth outcomes^36^. WQS regression follows a two-step process. First, we constructed a weighted index of mixture components to identify those most strongly associated with the outcome in either the positive or negative direction. Second, we identified the association between this weighted index and study outcomes (i.e., the overall mixture effect). In this study, we implemented WQS models using a decile transformation with 1,000 bootstraps, applying the same dataset for both training and validation to determine PAH index weights and evaluate OH-PAH mixture effects on BW and GA. Models were adjusted for key covariates as previously described. If statistically significant associations were observed between the WQS index and birth outcomes, we pre-specified the use of a permutation test to evaluate Type I error.^7,37^

The following sensitivity analyses were performed for the primary outcomes of BW and gestational ag: 1) exclusion of participants with maternal self-report of smoking tobacco and/or marijuana in the 24-hours prior to urine analysis (N=5) or maternal urinary cotinine concentration > 200 ng/mL (N=5); 2) exclusion of participants living with HIV (N=24) due to potentially different etiology of preterm birth 3) performing models with SG-adjusted urinary OH-PAH values, and 4) use of reported GA at delivery instead of calculated GA based on LMP. Analyses were performed using R version 4.4.1^38^ and the package gWQS.^39^

## 3. Results

### 3.1 Study population characteristics

There were N=400 pregnant women enrolled in ABC. Of these, n=387 delivered live births. Five participants were excluded due to multiple gestation, and two were excluded due to missing information on BW or GA. Nineteen participants did not provide a usable third trimester urine sample and 8 urine samples were lost due to spills/breakage or contamination. The resulting study population comprised n=353 participants. Infants in the study population were 52.1% male (Table 1). Mothers had a median age of 26.7 years (interquartile range [IQR]: 23.4-32.6 years) at delivery. Most mothers completed secondary school (56.7%), while 27.2% completed primary school or less and 16.1% attended college, university, or vocational school. The majority (55.8%) identified as homemakers, while 37.7% reported waged employment. The prevalence of flush toilets among the study participants was 96.9%, with the remainder using pit latrines. Over half of participants (55.2%) reported having a one-room home, while 31.7% lived in two-room homes and 13.0% had three or more rooms. About a third (33.7%) reported exposure to smoke from dirty fuel (charcoal, wood, kerosene, brickettes) every day, 26.3% said some days, and 39.9% said rarely or not at all. Among the most common sources of indoor combustion exposure were burning of rubbish and mosquito repellant (30.2% and 17.2%, respectively, reporting exposure daily, most days, or some days; Table S1). The most common daily exposures to outdoor combustion sources were cooking smoke (51.6%), vehicle smoke (50.1%), and dumpsite-related burning (22.5%). The mean BW in the study sample was 3.2 kg (SD: 0.45 kg) and the mean GA was 276 days (39.4 weeks) (Table 2). The prevalence of preterm birth in the study sample was 7.6%. Maternal urinary cotinine during pregnancy was < 10 ng/mL in 96.6% of the study sample; 2% had urinary cotinine 10-200 ng/mL and 1.4% had urinary cotinine > 200 ng/mL.

**Table 1.** Descriptive characteristics of pregnant participants at study enrollment (N=353)

| Characteristic | Frequency or Median | Percent or IQR |
| --- | --- | --- |
| **Maternal age at delivery** |  |  |
| Median (IQR) | 26.7 | 9.2 |
| **Maternal education** |  |  |
| Primary, pre-primary, or one | 96 | 27.2% |
| Secondary | 200 | 56.7% |
| College/University/Vocational | 57 | 16.1% |
| **Maternal marital status** |  |  |
| Divorced/separated/widowed | 12 | 3.4% |
| Married | 303 | 85.8% |
| Never married/single | 38 | 10.8% |
| **Maternal employment status** |  |  |
| Homemaker | 220 | 62.3% |
| Employed | 133 | 37.7% |
| **Parity** |  |  |
| Zero | 84 | 23.8% |
| One | 120 | 34.0% |
| Two | 82 | 23.2% |
| Three or more | 67 | 19.0% |
| **Pollutant cook fuel use** |  |  |
| Daily or most days | 119 | 33.7% |
| Some days | 93 | 26.3% |
| Rarely or not at all | 141 | 39.9% |
| **Toilet type** |  |  |
| Flush toilet | 342 | 96.9% |
| Pit latrine | 11 | 3.1% |
| **Number of rooms in house** |  |  |
| One | 195 | 55.2% |
| Two | 112 | 31.7% |
| Three or more | 46 | 13.0% |
| **Urinary specific gravity** |  |  |
| Median (IQR) | 1.015 | 0.010 |
| **Urinary cotinine (ng/mL)** |  |  |
| Median (IQR) |  |  |
| **Urinary cotinine category** |  |  |
| < 10 ng/mL | 341 | 96.6% |
| 10-200 ng/mL | 7 | 2.0% |
| >200 ng/mL | 5 | 1.4% |

**Table 2.**
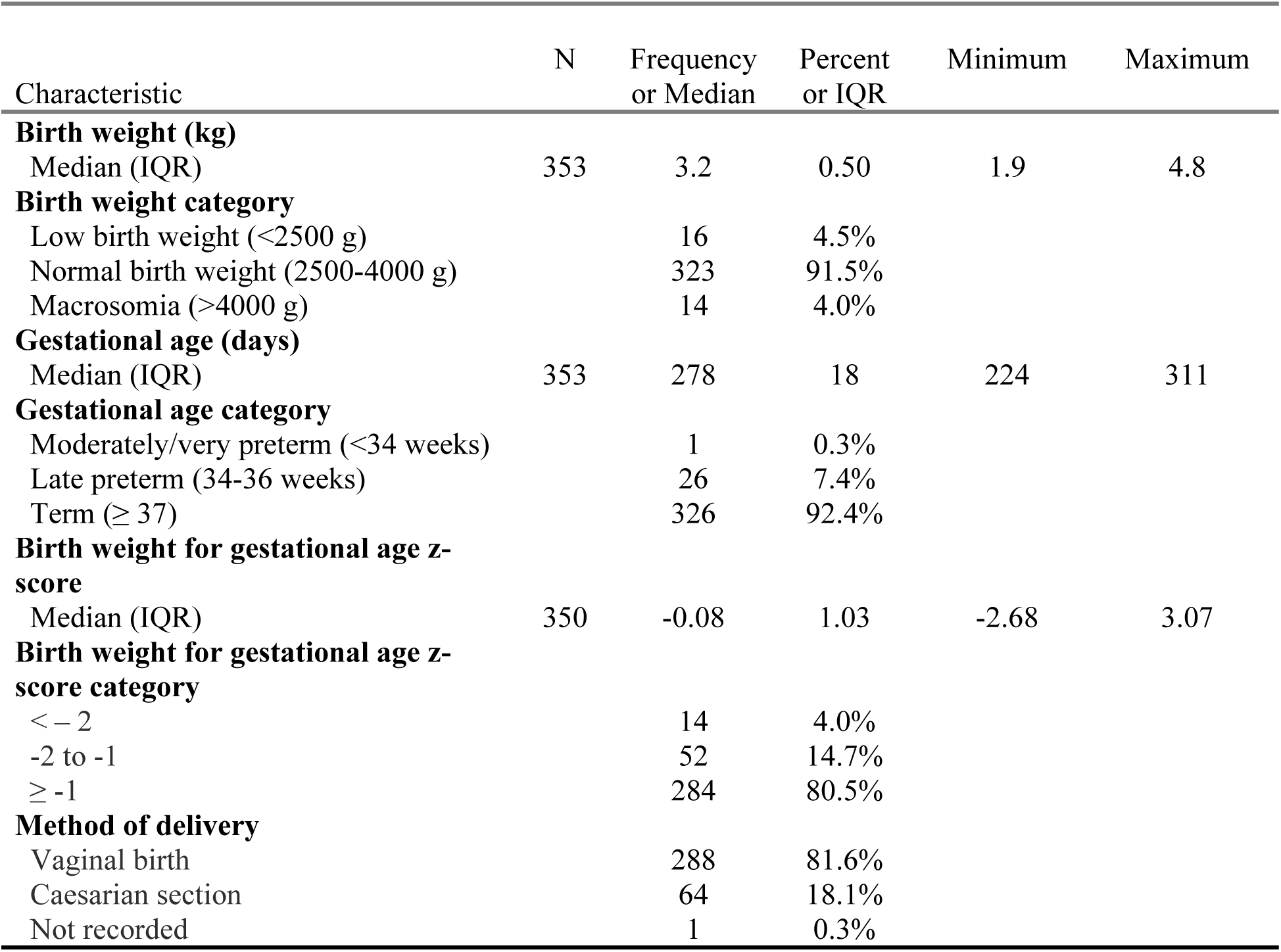
Birth outcomes in the study population.

### 3.2 Pregnancy OH-PAH exposure

Some analytes were not quantifiable in certain samples due to interfering peaks. This affected n=4 2/9- OH-FLUO, n=5 2-OH-PHEN, and n=6 3-OH-PHEN measurements; these measurements were imputed with the median of the analyte distribution. Due to matrix interference from fluorescent material in the urine that co-elutes with 1-OH-NAP, quantification of this compound was unreliable; 1-OH-NAP was therefore excluded as an analyte in the present analysis.

Detection frequencies for the five included OH-PAH metabolites ranged from 99.1% to 100% (Table 3). The geometric mean concentrations concentration ranged from 0.16 ng/mL (3-OH-PHEN) to 17.5 ng/mL (2-OH-NAP). 2-OH-NAP was weakly to moderately correlated with other metabolites (Pearson correlation coefficient [r]: 0.28-0.43; Figure S1). Other metabolites were moderately to strongly correlated (r: 0.60-0.75).

**Table 3.** Maternal urinary mono-hydroxylated polycyclic aromatic hydrocarbon (OH-PAHs) levels in ng/mL (n=353)

| OH-PAH | Arithmetic Mean | Geometric mean | SD | IQR | P25 | Median | P75 | Max | LOD | Detection rate |
| --- | --- | --- | --- | --- | --- | --- | --- | --- | --- | --- |
| 2-OH-NAP | 28.5 | 17.5 | 38.2 | 22.6 | 10.3 | 18.4 | 32.9 | 457.7 | 3.52 | 100% |
| 2/9-OH-FLUO | 0.85 | 0.54 | 0.96 | 0.66 | 0.31 | 0.57 | 0.97 | 8.01 | 0.013 | 99.1% |
| 2-OH-PHEN | 0.45 | 0.32 | 0.41 | 0.41 | 0.20 | 0.32 | 0.61 | 2.82 | 0.002 | 100% |
| 3-OH-PHEN | 0.24 | 0.16 | 0.27 | 0.19 | 0.09 | 0.16 | 0.28 | 2.03 | 0.002 | 100% |
| 1-OH-PYR | 0.86 | 0.52 | 1.21 | 0.60 | 0.29 | 0.52 | 0.89 | 13.20 | 0.011 | 99.7% |

Abbreviations: 2-OH-NAP – 2-hydroxynaphthalene; 2/9 FLUO – combined 2-and 9- hydroxyfluorene; 2-OH-PHEN – 2-hydroxyphenanthrene; 3-OH-PHEN – 3-hydroxyphenanthrene; 1-OH-PYR – 1-hydroxypyrene.

Infant sex and maternal age, education, marital status, and employment status were not markedly different between participants in different tertiles of OH-PAH molar sums (Table S3). More participants in the second and third tertiles of OH-PAH exposure had urinary cotinine levels > 10 ng/mL (4.2%) relative to participants in the lowest tertile (1.7%). More participants in the third tertile of OH-PAH exposure also reported daily exposure to dirty cooking fuels (54.7%) relative to those in the second tertile (31.4%) and first tertile (15.3%).

### 3.3 Analytic results

The association between individual OH-PAHs and BW was null in primary and co-exposure models (Figure 1, Table S4). With primary model adjustment, there was a statistically significant negative association between 2-OH-PHEN and GA, with a decrease of 1.6 days associated with a doubling in 2- OH-PHEN concentrations (95% confidence interval [CI]: -3.1, -0.1). With adjustment for co-exposure, this association was somewhat attenuated (beta coefficient [β] = -1.3, 95% CI: -2.9, 0.2). There was also a positive but not statistically significant association between 2-OH-PHEN and the secondary outcome of preterm birth (RR = 1.50 per doubling in 2-OH-PHEN concentration, 95% CI: 0.94, 2.38). These findings were largely unchanged in sensitivity analyses including exclusion of participants with positive HIV status, exclusion of participants with evidence of prenatal smoking, use of SG-adjusted OH-PAH values, and use of reported instead of calculated GA (Figure S2). OH-PAH concentrations were not associated with the secondary outcomes of BW-GA z-scores, though there was a positive association between 1-OH-PYR and BW-GA z-scores that approached statistical significance (β = 0.11, 95% CI: -0.01, 0.23).

**Figure 1.**
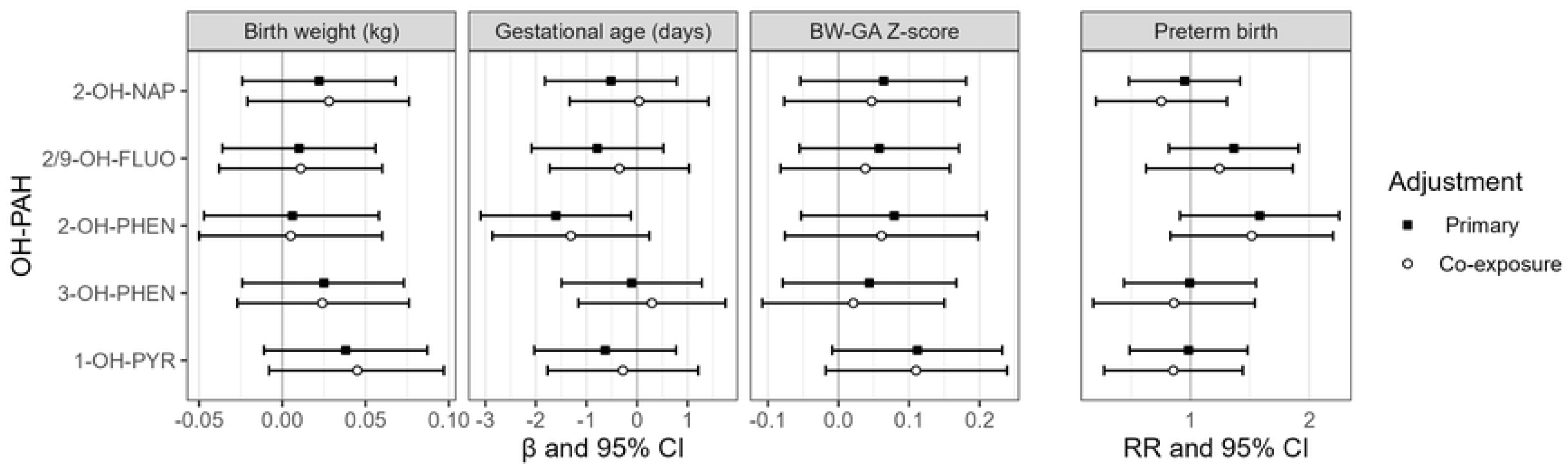
Estimates and 95 % confidence intervals (CI) for birth outcomes associated with a twofold increase in individual OH-PAH metabolites. Primary models were adjusted for urinary SG and maternal age, employment, education, marital status, and urinary cotinine category. Co-exposure models were additionally adjusted for use of dirty cooking fuel and exposure to indoor, outdoor, and work-related combustion sources. Abbreviations: 2-OH-NAP – 2-hydroxynaphthalene; 2/9 FLUO – combined 2-and 9-hydroxyfluorene; 2-OH-PHEN – 2-hydroxyphenanthrene; 3-OH-PHEN – 3-hydroxyphenanthrene; 1-OH-PYR – 1-hydroxypyrene.

In effect modification analyses, there were no consistent differences between effect estimates among male and female infants for the outcomes of BW and BW-GA z-scores (Figure 2). For the outcome of GA, effect estimates were consistently lower for female infants relative to male infants, with a significant interaction *p*-value for 3-OH-PHEN (*p* = 0.04). There was also an adverse association between 2-OH- PHEN and GA among female infants (β = -2.0, 95% CI: -3.8, -0.1) but not male infants (β = -0.2, 95% CI: -2.5, 2.1), though the interaction *p*-value was not significant (*p* = 0.19). For the outcome of preterm birth, effect estimates were also stronger among female infants, though without evidence of significant effect modification.

**Figure 2.**
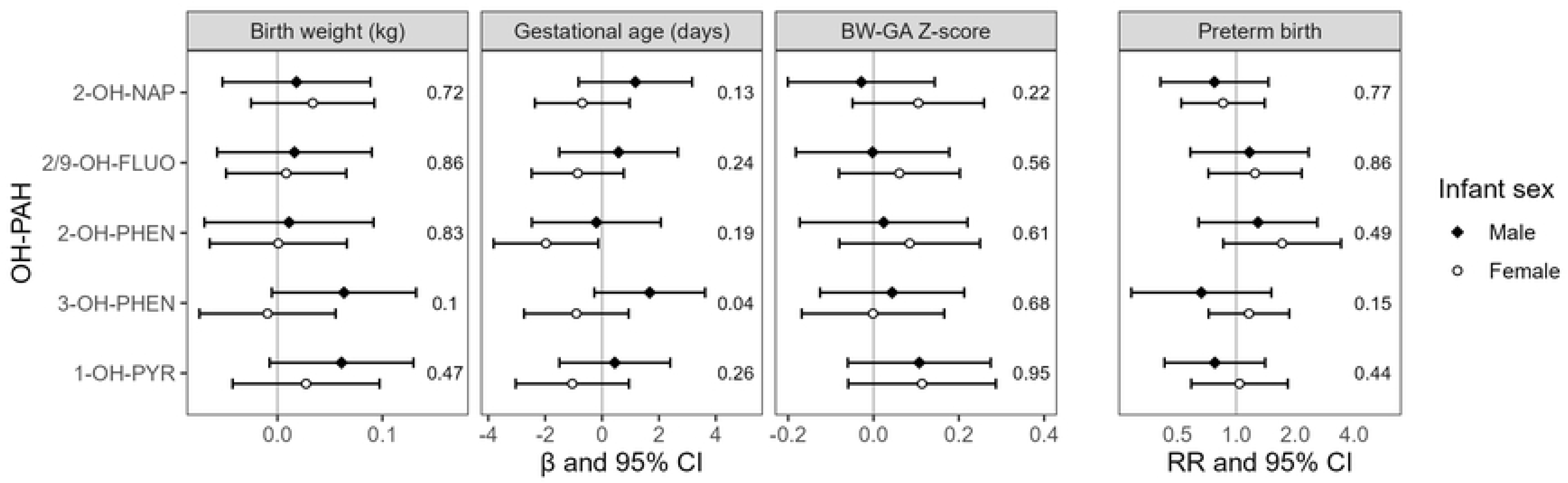
Analysis of effect modification by child sex determined from interaction models, including sex-specific effect estimates and interaction p-values. Models were adjusted for urinary SG and maternal age, employment, education, marital status, and urinary cotinine category. Abbreviations: 2-OH-NAP – 2-hydroxynaphthalene; 2/9 FLUO – combined 2-and 9- hydroxyfluorene; 2-OH-PHEN – 2-hydroxyphenanthrene; 3-OH-PHEN – 3-hydroxyphenanthrene; 1-OH-PYR – 1-hydroxypyrene.

In WQS models, associations between the WQS index (i.e., the overall OH-PAH mixture) and BW were null in both the positive and negative direction (Table S5). There was a non-statistically significant negative (adverse) association between the OH-PAH mixture and GA in primary models, with an effect estimate of -0.44 days per decile increase in the WQS Index, (95% CI: -0.98, 0.10) (Table 4). This association was somewhat attenuated in with co-exposure adjustment (β = -0.32, 95% CI: -0.88, 0.23). 2- OH-PHEN represented the largest contribution to the WQS index mixture representing the negative impact of the OH-PAH mixture on GA (Table 4). There was no evidence of a positive association between the OH-PAH mixture on GA (Table S5).

**Table 4.**
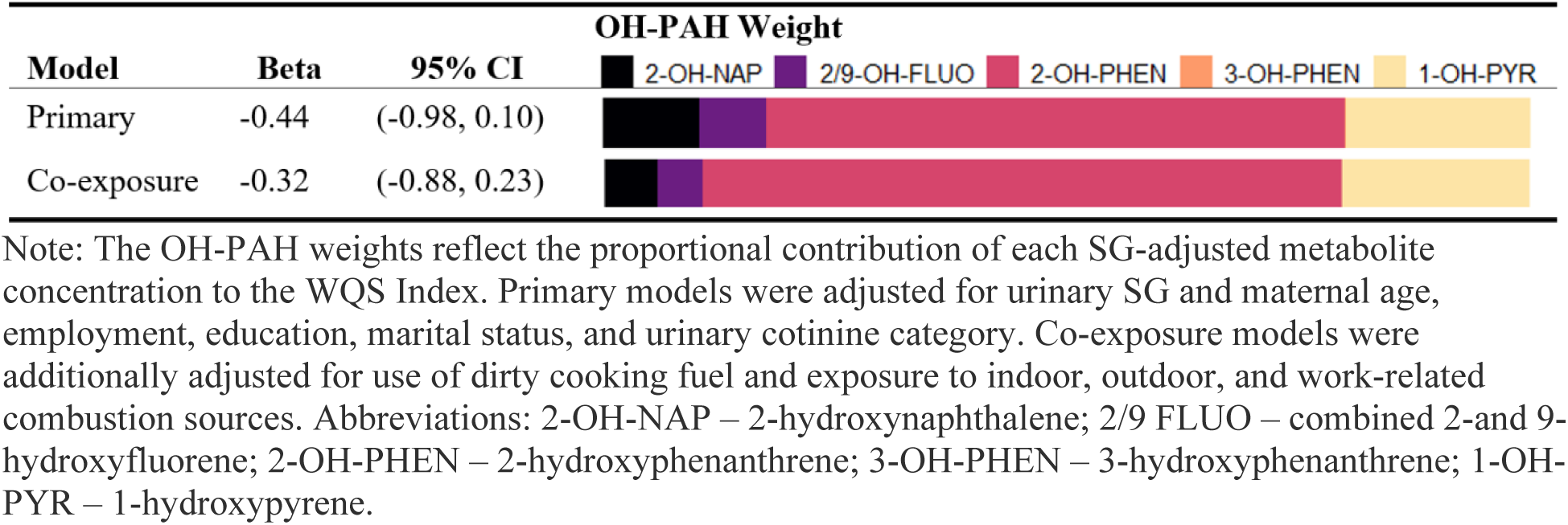
Association between the OH-PAH mixtures and gestational age (days) determined by weighted quantile sum (WQS) regression models in the negative direction.

## 4. Discussion

We explored associations between prenatal exposure to LMW PAHs and adverse birth outcomes in a prospective cohort in Nairobi, Kenya. We did not find evidence of associations between third trimester urinary OH-PAH concentrations and the outcomes of BW, BW-GA z-scores, or preterm birth in our study population. There was a significant association between 2-OH-PHEN and GA, which was stronger among female infants. There was also evidence of effect modification in the association between 3-OH-PHEN and GA, with a stronger adverse effect in female infants. We did not find evidence of an overall mixture effect of five OH-PAHs on either BW or GA.

Our study was conducted in a densely populated low-income urban settlement within close proximity to the Nairobi City dumpsite, thereby providing unique evidence from a sub-Saharan African low-income setting. The sources of PAHs in the study area differ from those in HICs. For example, indoor combustion of biomass cooking fuel sources has been shown to contribute to indoor concentrations of LMW PAHs in.^40,41^ While levels of ambient LMW PAHs have not been systematically evaluated in the study area, some HMW PAHs in Nairobi have been found to exceed the guideline levels established by the World Health Organization (WHO). For example, levels of ambient benzo[a]pyrene have been recorded at over 10 ng/m^3^, compared to the WHO guideline of 0.1 ng/m_3_, attributed to sources including industrial activities, motor vehicle traffic, and household biomass burning.^23,40,42^ The study site is also adjacent to the Dandora dump site, a recognized source of environmental pollution where burning of solid waste is common.^43–45^ While ambient PAH levels in the Dandora neighborhoods have not been systematically evaluated, LMW PAHs have been documented in soil, water, and air at other dump sites in Africa.^11,46^ Urinary PAH biomarkers in our study population reflect higher exposure relative to HIC settings; for example, the SG-adjusted geometric mean concentration of 2-OH-NAP in the ABC Cohort was over five times greater than in the multi-site U.S. ECHO PATHWAYS pregnancy cohort (17.5 vs 3.61 ng/mL); the mean concentration of 1-OH-PYR was over seven times greater (0.52 vs 0.07 ng/mL). Therefore, our analysis explores exposure-response relationships in a higher range of urinary OH-PAH concentrations and in a setting with a unique exposure profile.

Prior studies have evaluated associations between maternal PAHs and birth outcomes, though the findings have been inconsistent.^16–18^ A recent systematic review identified 31 studies evaluating associations between PAH exposure and birth outcomes, including 15 cohort studies.^17^ These studies have largely been conducted in HICs, as well as China and Poland, and have used a variety of exposure assessments methodologies, study designs, and outcome definitions. While some studies have also evaluated urinary biomarkers of exposure in pregnancy cohorts, ^19,47–51^ others have used personal air monitoring, ambient air monitoring, dietary estimates from food frequency questionnaires, and levels of PAHs or DNA adducts in blood or placenta to quantify exposure.^17,18,52–54^ Among those that have evaluated urinary OH-PAH metabolites, differences in the specific analytes and pregnancy timepoints measured can also complicate comparison between studies and with the present analysis. Freije et al.^19^ explored associations between maternal mid-pregnancy urinary OH-PAHs and GA in 1,677 participants of the multi-site ECHO PATHWAYS prospective cohort in the U.S.—among the largest cohort studies using urinary biomarkers of exposure. The authors identified a small but significant association between maternal urinary 2-OH- NAP and decreased GA at birth (-1.92 days per 10-fold higher concentration, 95% CI: -3.13, -0.70). In contrast to our findings, no association was found for 2-OH-PHEN. Yang et al.^47^ also evaluated maternal urinary OH-PAHs and GA in a cohort study in China (N=106). A borderline negative association was observed between 2-OH-PHEN measured in the second trimester and GA, but not for 2-OH-PHEN in the first or third trimester, or other OH-PAH metabolites. In a nested case-control study of 458 births in the U.S. LIFECODES cohort, combined urinary 2- and 3-OH-PHEN measured at various time points in pregnancy showed an inverse but nonsignificant association with preterm birth.^48^ A systematic review found that three out of ten studies of prenatal PAHs and GA identified significant adverse associations, but differences in study design and exposure assessment precluded meta-analysis of results.^17^

Our analysis did not find associations between OH-PAH metabolites and BW. Among 19 studies included in a recent systematic review, ten studies (including seven cohort studies) identified significant associations between PAH exposure and BW, while nine studies found no association. A meta-analysis of four studies evaluating the association between maternal urinary 1-OH-PYR and term BW also found no association (OR = 1.00 per log ng/g creatinine increase, 95% CI: 0.97, 1.03).^16^ While fewer studies have evaluated the outcome of BW-GA z-scores, multiple metabolites collected throughout pregnancy were associated with reduced BW-GA z-scores in the U.S. LIFECODES cohort (N=458). The strongest effect estimate was observed for 2-OH-NAP (β = -0.22 per IQR increase, 95% CI: -0.36, -0.08).^48^

Several biological mechanisms may underly associations between PAH exposure and adverse birth outcomes, including oxidative stress, systemic inflammation, endocrine disruption, and other epigenetic impacts. Human and toxicological studies have found that PAH exposure during pregnancy leads to increased levels of inflammatory and oxidative stress biomarkers.^55,56^ For example, third-trimester exposure to certain LMW PAHs was associated with increased plasma inflammation marker (C-reactive protein) and oxidative stress biomarkers 8-isoprostane and 8-hydroxydeoxyguanosine.^56^ Both oxidative stress and systemic inflammation have been implicated as important contributors to preterm birth, including spontaneous preterm birth, as well as placental dysfunction and impaired fetal development.^19,57–61^ Prenatal PAH exposure has also been associated with changes in hormonal regulation among pregnant women, including increased concentrations of corticotropin-releasing hormone (CRH) concentrations and decreased ratio of progesterone to estriol.^21^ Elevated CRH and progesterone withdrawal are both involved in the initiation of labor and implicated in the etiology of preterm birth.^62,63^ Finally, PAH exposure has been linked to epigenetic changes in placental gene expression, including genes associated with immune function and vitamin absorption, that could impact fetal development and perinatal outcomes.^20,54,64^ Factors influencing maternal susceptibility such as nutritional deficiencies, pre-existing health conditions (including HIV infection), and co-exposures to other pollutants may exacerbate the impact of PAH exposure on these biological pathways and resulting adverse birth outcomes.^65–67^

We found some evidence for effect modification by infant sex, with a significant association between 2- OH-PHEN and GA among participants with female infants only. However, the interaction p-value for this association was not statistically significant. There was evidence of effect modification in the association between 3-OH-PHEN and GA. In the multi-site PATHWAYS prospective cohort study, Freije et al.^19^ also identified evidence of effect modification by infant sex for the association between 2-OH-PHEN and preterm birth. Similar to the present analysis, a stronger adverse effect was estimated for female infants. However, contrary to our findings, the authors also found evidence of effect modification by infant sex in the association between 1-OH-PYR and preterm birth. There is some evidence for sex-dimorphic biologic impacts of PAHs during pregnancy. Paquette et al.^20^ found that the impact of OH-PAHs on the placental transcriptome varied with child sex, with OH-PAHs associated with the placental expression of 60% more genes in female births relative to male births—including genes related to vitamin absorption. Cathey et al.^21^ also identified associations between certain OH-PAHs and CRH, progesterone, estriol, and testosterone among female infants but not male infants. Additional research would clarify the potential role of fetal sex in modifying the biologic impacts of specific OH-PAHs and downstream effects on pregnancy outcomes.

Other environmental or social determinants of health may co-occur with PAH exposure and have independent or synergistic effects on pregnancy outcomes. We aimed to control for confounding by combustion-generated co-pollutants using questionnaire-based assessment of indoors, outdoors, and work-related combustion sources. While the findings were not significantly different in this ‘co-exposure’ model, the significant association between 2-OH-PHEN and GA was attenuated with additional adjustment. This may suggest that some of the observed association in the primary model could be attributed to the role of other pollutants generated by combustion that are independently associated with birth outcomes. Additional research exploring the impact of co-exposure to multiple pollutants could improve understanding of the biologic mechanisms of adverse health outcomes and role of exposure mixtures.

Our study has several strengths. We add to a limited number of studies evaluating PAHs and birth outcomes using a prospective cohort design and maternal urinary biomarkers of exposure.^19,47–51^ We focused on an understudied LMIC study population and achieved high retention, minimizing potential selection bias, retaining a larger study population than comparable HIC studies.^47,50,51^ We also paired biomarkers of exposure with rich questionnaire-based evaluation of potential co-exposure to indoor, outdoor, and occupational combustion sources. This framework offers unique opportunity to evaluate the influence of PAHs on birth outcomes in Sub-Saharan Africa, and will serve as a framework to explore health impacts throughout early life.

Our analysis also has limitations. First, PAH exposure assessment was based on a single spot urine sample collected during the third trimester, which may not accurately reflect typical exposure levels over the course of pregnancy. Urinary OH-PAHs have relatively short biological half-lives—generally between 2.5 and 12 hours—and therefore primarily indicate recent exposure.^68,69^ Research examining urinary OH-PAH concentrations throughout pregnancy has reported intraclass correlation coefficients (ICCs) that vary considerably by specific analyte and population subgroup.^6,70–72^ Given this variability, use of a single measurement may introduce exposure misclassification and reducing our ability to detect true associations with birth outcomes. Additionally, our analysis focused solely on metabolites of LMW PAHs, omitting high molecular weight PAHs that are primarily excreted via feces but which may also be relevant to pregnancy outcomes.^73^ Imperfect recall or recall bias may also affect the accuracy of reported LMP and therefore our calculated GA measure; however, we found that our findings were largely unchanged when using clinician-reported GA as an outcome in place of calculated GA. There may also be residual confounding from covariates that were either not measured or measured with limited precision. We used questionnaire responses to evaluate exposure to combustion sources that may influence birth outcomes through generation of co-pollutants besides PAHs; these responses introduce the potential for recall bias and/or exposure misclassification from imperfect recall. While our unique study setting is a strength, the findings may not be generalizable to rural populations or regions with different sources and levels of PAH exposure. Future multi-site studies in rural and urban LMIC contexts could provide broader insights into regional variations in PAH exposure, health impacts, and potential intervention strategies.

This study is among the first to evaluate PAHs and adverse birth outcomes in an urban LMIC setting. We did not identify strong evidence of associations between OH-PAHs and birth outcomes, though there was evidence of an impact of 2-OH-PHEN on GA, with a stronger association among female infants. Given the widespread sources of indoor and ambient PAHs in Nairobi and other LMIC contexts, evaluating potential health impacts and preventing exposure are paramount. In particular, policies aimed at reducing air pollution, improving access to household electricity, and incorporating environmental health into prenatal care could help mitigate the impact of PAHs on maternal and infant health. Additional research on the health effects of PAHs and intervention strategies would inform targeted interventions to prevent exposure.

## Data Availability

Data are available in the supporting information material.

## Acknowledgements

We are grateful for the participation of families enrolled in the ABC cohort, as well as the dedication of research staff and investigators. In particular, we thank the ABC study field team, including Emily Adhiambo, Brendah Isavwah, Vincent Kipter, James Lele, Edith Lumumba, Bernard Makau, Margaret Muruthi, Charity Muthui, Laura Mwangi, Perpetual Nyaguthi, and Electine Oyuga. The ABC study was funded by U.S. NIEHS R01ES032153 to SBN and University of Washington EDGE Center NIEHS P30ES007033. The content is solely the responsibility of the authors and does not necessarily represent the official views of the National Institutes of Health.

